# Evaluation of the diagnostic value of YiDiXie^™^-SS, YiDiXie^™^-HS and YiDiXie^™^-D in prostate cancer

**DOI:** 10.1101/2024.06.26.24309554

**Authors:** Xutai Li, Zhenjian Ge, Qingshan Yang, Chen Sun, Wenkang Chen, Yingqi Li, Shengjie Lin, Pengwu Zhang, Wuping Wang, Siwei Chen, Yutong Wu, Huimei Zhou, Wei Li, Rong Huang, Liangchao Ni, Yongqing Lai

**Author notes:** Corresponding author: Yongqing Lai, Peking University Shenzhen Hospital, 1120 Lianhua Road, Shenzhen 518036, E mail; Liangchao Ni, Peking University Shenzhen Hospital, 1120 Lianhua Road, Shenzhen 518036, E mail; Rong Huang, Peking University Shenzhen Hospital, 1120 Lianhua Road, Shenzhen 518036, E mail. Contributed equally to this work (Xutai Li, Zhenjian Ge, Qingshan Yang).

## Abstract

**Background:** Prostate cancer is one of the most common malignant tumors and poses a substantial threat to human health. PSA or enhanced MRI is widely used in prostate cancer screening or diagnosis. However, false-positive PSA or enhanced MRI results can lead to misdiagnosis and incorrect puncture biopsy, while false-negative PSA or enhanced MRI results can lead to missed diagnosis and delayed treatment. There is an urgent need to find convenient, economical and non-invasive diagnostic methods to reduce the PSA or enhanced MRI false-negative and false-positive rates. The aim of this study was to evaluate the diagnostic value of YiDiXie™-SS, YiDiXie™-HS and YiDiXie™-D in prostate cancer.

**Patients and methods:** The study finally included 464 subjects with positive PSA test (prostate cancer group, n=292; BPH group, n=172). Remaining serum samples from the subjects were collected and tested with YiDiXie™ all-cancer detection kit. The sensitivity and specificity of YiDiXie™-SS, YiDiXie™-HS and YiDiXie™-D were evaluated respectively.

**Results:** The sensitivity of YiDiXie™-SS was 100% (95% CI: 98.7% - 100%) and its specificity was 60.5% (95% CI: 53.0% - 67.5%). This means that YiDiXie™-SS has an extremely high sensitivity and relatively high specificity in prostate tumors.YiDiXie™-HS has a sensitivity of 93.8% (95% CI: 90.5% - 96.1%) and a specificity of 86.0% (95% CI: 80.1% - 90.4%). This means that YiDiXie™ -HS has high sensitivity and specificity in prostate tumors.YiDiXie™-D has a sensitivity of 80.8% (95% CI: 75.9% - 84.9%) and a specificity of 93.0% (95% CI: 88.2% - 96.0%). This means that YiDiXie™-D has relatively high sensitivity and very high specificity in prostate tumors.YiDiXie™-SS had a sensitivity of 100% (95% CI: 98.6% - 100%), 100% (95% CI: 97.6% - 100%) and specificity of 57.7% (95% CI: 49.8% - 65.2%), 59.6% (95% CI. 49.5% - 68.9%). This means that the application of YiDiXie™-SS reduced the false positive rate of PSA and enhanced MRI by 57.7% (95% CI: 49.8% - 65.2%), 59.6% (95% CI: 49.5% - 68.9%), respectively, with essentially no increase in the underdiagnosis of malignant tumors.YiDiXie™-HS had a sensitivity of 92.3% (95% CI: 66.7% - 99.6%), 90.2% (95% CI: 79.0% - 95.7%), and specificity of 81.3% (95% CI: 57.0% - 93.4%), 83.3% (95% CI : 55.2% - 97.0%),respectively. This means that the application of YiDiXie™-HS reduced the false-negative rate of PSA and enhanced MRI by 92.3% (95% CI: 66.7% - 99.6%), 90.2% (95% CI: 79.0% - 95.7%), respectively.The sensitivity of YiDiXie™-D in PSA and enhanced MRI positive patients was 81.0% (95% CI: 76.0% - 85.2%), 83.6% (95% CI: 77.1% - 88.6%), respectively, and its specificity was 92.9% (95% CI: 87.8% - 96.0%), 92.6% (95% CI : 85.4% - 96.3%), respectively. This means that YiDiXie™-D reduced PSA and enhanced MRI false positive rates by 92.9% (95% CI: 87.8% - 96.0%), 92.6% (95% CI: 85.4% - 96.3%), respectively.YiDiXie™-D had a sensitivity of 76.9% (95% CI: 49.7% - 91.8%) and 76.5% (95% CI: 63.2% - 86.0%) in PSA- and enhanced MRI-negative patients, respectively, and its specificity was 93.8% (95% CI: 71.7% - 99.7%) and 83.3% (95% CI : 55.2% - 97.0%), respectively. This means that YiDiXie™-D reduced PSA and enhanced MRI false-negative rates by 76.9% (95% CI: 49.7% - 91.8%) and 76.5% (95% CI: 63.2% - 86.0%), respectively, while maintaining a high specificity..

**Conclusion:** YiDiXie™-SS has very high sensitivity and relatively high specificity in prostate tumors.YiDiXie™-HS has high sensitivity and high specificity in prostate tumors.YiDiXie ™ -D has relatively high sensitivity and very high specificity in prostate tumors. YiDiXie ™-SS significantly reduces the rate of PSA or enhanced MRI false positives with essentially no increase in delayed treatment for prostate cancer.YiDiXie™-HS significantly reduces the rate of PSA or enhanced MRI false negatives.YiDiXie™-D significantly reduces the rate of PSA or enhanced MRI false positives or significantly reduces the rate of false negatives thereof while maintaining a high level of specificity. YiDiXie™ tests can play an important role in prostate cancer, and are expected to solve the problem of “high false-positive rate” and “high false-negative rate” of PSA or enhanced MRI.

**Clinical trial number:** ChiCTR2200066840.

## INTRODUCTION

Prostate cancer is one of the most common malignant tumors. According to the latest data for 2022, there are 1.5 million new cases of prostate cancer and 397,000 new deaths worldwide, making it the second most prevalent malignancy and the fifth most common cause of cancer-related deaths among males^1^. Compared with 2020, prostate cancer incidence and death rates in 2022 increased by 3.71% and 5.72%, respectively^1,2^. When discovered and diagnosed early screening, the survival rate for individuals with localized prostate cancer can be as high as 99% at 10 years or more; However, the overall 5-year survival rate for men diagnosed with advanced disease (distant metastases) is only about 30%^3,4^. Therefore, prostate cancer poses a significant risk to human health.

PSA and enhanced MRI is widely used in prostate cancer screening and diagnosis. On one hand, PSA and enhanced MRI can produce a large number of false-positive results^5-8^. The PLCO trial indicated that among men who underwent prostate biopsy following a positive screening, only 32.3% were diagnosed with prostate cancer within 120 days^8^. In another study, among all screened men who underwent biopsy after a positive screening, only 24.7% were diagnosed with prostate cancer within 12 months of testing^9^. In another study, only 24.7% of all screen-positive men who underwent biopsy were diagnosed with prostate cancer within 12 months after testing^9^. When the PSA or enhanced MRI test is positive, patients usually undergo a puncture biopsy for further diagnosis^10^. False-positive results for prostate cancer mean that patients will have to undergo puncture biopsy for further diagnosis, and therefore these patients will have to bear unnecessary mental suffering, expensive tests, physical injuries and other negative consequences^11,12^. Hence, there is an urgent need to establish a convenient, affordable, and non-invasive diagnostic approach to lower the false-positive rate of PSA and enhanced MRI for prostate cancer.

On the other hand, PSA and enhanced MRI can produce a large number of false-negative results^5-8^.When PSA or enhanced MRI is negative, patients are usually taken for observation, with regular follow-up^10^. False-negative PSA or enhanced MRI results imply a missed diagnosis of prostate cancer, which will likely lead to delayed treatment, progression of malignancy, and possibly even development of advanced stages. Patients will thus have to bear the adverse consequences of poor prognosis, high treatment costs, poor quality of life, and short survival. Therefore, there is an urgent need to find a convenient, economical and noninvasive diagnostic method to reduce the false-negative rate of PSA and enhanced MRI.

Based on the detection of miRNAs in serum, Shenzhen KeRuiDa Health Technology Co., Ltd. has developed “YiDiXie ™ all-cancer test” (hereinafter referred to as YiDiXie ™ tests)^13^. With only 200 milliliters of whole blood or 100 milliliters of serum, the test can detect multiple cancer types, enabling detection of cancer at home^13^. YiDiXie ™ tests consists of three independent tests: YiDiXie ™-HS, YiDiXie™-SS and YiDiXie™-D^13^.

The purpose of this study was to evaluate the diagnostic value of YiDiXie™-SS, YiDiXie™-HS and YiDiXie™-D in prostate cancer.

## PATIENTS AND METHODS

### Study design

This work is part of the sub-study “Evaluating the diagnostic value of YiDiXie™ tests in multiple tumors”of the SZ-PILOT study (ChiCTR2200066840).

The SZ-PILOT study (ChiCTR2200066840) was a single-center, prospective, observational study. Subjects who signed the broad informed consent for donation of remaining samples at the time of admission or medical health checkup were included, and 0.5 ml of their remaining serum samples were collected for this study.

This study was blinded. Neither the laboratory personnel performing YiDiXie ™ tests nor the technicians of KeRuiDa Co. evaluating the raw results of YiDiXie ™ tests were informed of the subject’s clinical information. The clinical experts assessing the subjects’ clinical information were also unaware of the results of YiDiXie™ tests.

The study was approved by the Ethics Committee of Peking University Shenzhen Hospital and was conducted in accordance with the International Conference on Harmonization for “Good clinical practice guidelines” and the Declaration of Helsinki.

### Participants

Subjects in the two groups were enrolled separately, and all subjects who met the inclusion criteria were included consecutively.

The malignant group initially enrolled hospitalized patients with “suspected (solid or hematological) malignant tumors” with a signed broad informed consent for donation of the remaining samples. Subjects with a pathologic diagnosis of “prostate cancer” were included in the malignant group, and those with a pathologic diagnosis of “benign prostatic hyperplasia” were included in the benign group. Participants who had ambiguous pathologic results were excluded from the study. Some of these prostate cancer samples were used in our prior work^13^.

Subjects who were not qualified in the serum sample quality test prior to YiDiXie ™ tests were excluded from the study. For further information on enrollment and exclusion, please see our prior work^13^.

### Sample collection, processing

The serum samples used in this study were obtained from serum left over after a normal consultation, without the need for additional blood sampling. Approximately 0.5 ml of serum was collected from the remaining serum of the participants in the Medical Laboratory and stored at - 80°C for use in the subsequent YiDiXie™ test.

### YiDiXie™ tests

YiDiXie™ tests is performed using the “YiDiXie ™ all-cancer detection kit”. The “YiDiXie ™ all-cancer detection kit” is an in-vitro diagnostic kit developed and manufactured by Shenzhen KeRuiDa Health Technology Co., Ltd. for use in fluorescent quantitative PCR instruments. It detects the expression levels of dozens of miRNA biomarkers in serum to determine whether cancer is present in the subject. It predefines appropriate thresholds for each miRNA biomarker, ensuring that each miRNA marker has a high specificity. The YiDiXie ™ kit integrates these independent assays in a concurrent testing model to significantly increase the sensitivity in broad-spectrum cancers and maintain a high specificity.

YiDiXie ™ tests consists of three tests with highly different characteristics: YiDiXie ™ -HS, YiDiXie ™ -SS and YiDiXie ™ -D. The YiDiXie ™ -HS (YiDiXie™-Highly Sensitive) is developed with high sensitivity and high specificity. YiDiXie™-SS (YiDiXie ™ -Super Sensitive) significantly increases the number of miRNA tests to achieve extremely high sensitivity for all stages in all malignancy types. YiDiXie ™ -D (YiDiXie ™ -Diagnosis) significantly increases the diagnostic threshold of individual miRNA tests to achieve very high specificity.

Perform YiDiXie ™ tests according to the instructions of the “YiDiXie™ all-cancer detection kit”. Refer to our prior work for detailed procedures^13^.

The original test results were analyzed by the laboratory technicians of KeRuiDa Co. and determined to be “positive” or “negative”.

### PSA test

The cutoff value for PSA test is 4ng/ml. TPSA ≥ 4ng/ml is considered “positive” for PSA test and TPSA < 4ng/ml is considered “negative”.

### Diagnosis of enhanced MRI

If there is a PI-RADS grade in the diagnosis of enhanced MRI of the prostate, a PI-RADS grade of 4 or 5 is regarded as “ positive “, and a PI-RADS grade of ≤3 is regarded as “negative”.

If there is no PI-RADS grade in the diagnosis of enhanced MRI of the prostate, the results of the examination are considered positive or negative according to the diagnostic conclusion. If the diagnostic conclusion is positive, more positive, or favors a malignant tumor, the result is considered positive. If the diagnosis is positive, more positive or inclined to benign disease, or if the diagnosis is ambiguous, the test result is judged as “negative”.

### Clinical data collection

Clinical, pathological, laboratory, and imaging data in this study were extracted from the subjects’ hospitalized medical records or physical examination reports. Clinical staging was completed by trained clinicians assessed according to the AJCC staging manual (7th or 8th edition)^14,15^.

### Statistical analyses

Descriptive statistics were presented for baseline attributes and demographic information. For continuous variables, the total number of subjects (n), mean, standard deviation (SD) or standard error (SE), median, first quartile (Q1), third quartile (Q3), minimum, and maximum values were calculated. For categorical variables, the number and percentage of subjects in each category were calculated. The Wilson (score) technique was used to compute the 95% confidence intervals (CI) for several indicators.

## RESULTS

### Participant disposition

464 study participants were involved in this research (n = 292 cases for the malignant group and 172 cases for the benign group). The 464 participants’ clinical and demographic details are listed in Table 1.

**Table 1.** Participants’ demographic and clinical manifestation.

|  | Malignant (n = 292) | Benign (n =172) | Total (N = 464) |
| --- | --- | --- | --- |
| Age, years |  |  |  |
| Mean (SD) | 68.7 (8.74) | 63.9 (8.61) | 66.9 (8.99) |
| Median (Q1,Q3) | 69 (63, 75) | 64 (59, 70) | 67 (61, 74) |
| Min, max | 44, 87 | 36, 88 | 36, 88 |
| Age, group, n (%) |  |  |  |
| <50 | 8 (2.7) | 8 (4.6) | 16 (3.4) |
| ≥50 | 284 (97.3) | 164 (95.3) | 448 (96.6) |
| <65 | 93 (31.8) | 96 (55.8) | 189 (40.7) |
| ≥65 | 199 (68.2) | 76 (44.2) | 275 (59.3) |
| Body mass index (kg/m2) |  |  |  |
| n | 187 | 141 | 328 |
| Mean (SD) | 23.6 (2.86) | 23.9 (2.91) | 23.7 (2.88) |
| Median (Q1,Q3) | 23.4 (21.8, 25.6) | 23.8 (22.0, 25.9) | 23.6 (21.9, 25.6) |
| Min, max | 16.5 31.2 | 15.6 32.0 | 15.6 32.0 |
| Body mass index category, n (%) |  |  |  |
| Underweight | 8 (2.7) | 3 (1.7) | 11 (2.4) |
| Benign | 96 (32.9) | 72 (41.9) | 168 (36.2) |
| Overweight | 73 (25.0) | 55 (32.0) | 128 (27.6) |
| Obese | 10 (3.4) | 11 (6.4) | 21 (4.5) |
| Missing | 105 (40.0) | 31 (18.0) | 136 (29.3) |
| TPSA value |  |  |  |
| (0, 4) | 13 (4.5) | 16 (9.3) | 29 (6.3) |
| [4, 10) | 79 (27.0) | 100 (58.1) | 179 (38.6) |
| [10, 20) | 71 (24.3) | 43 (25.0) | 114 (24.6) |
| [20, 100) | 76 (26.0) | 12 (7.0) | 88 (19.0) |
| [100, 1000+) | 53 (18.2) | 1 (0.6) | 54 (11.6) |
| AJCC clinical stage |  |  |  |
| Stage I | 6 (2.1) |  | 6 (2.1) |
| Stage II | 161 (55.1) |  | 161 (55.1) |
| Stage III | 15 (5.1) |  | 15 (5.1) |
| Stage IV | 109 (37.3) |  | 109 (37.3) |
| Missing | 1 (0.3) |  | 1 (0.3) |

Q1,Q3, first quartile, third quartile; SD, standard deviation.

In terms of clinical and demographic traits, the two study subject groups were similar (Table 1). The mean (standard deviation) age was 66.9 (8.99) years.

### Diagnostic Performance of YiDiXie™-SS

As shown in Table 2, the sensitivity of YiDiXie™ -SS was 100% (95% CI: 98.7% - 100%) and its specificity was 60.5% (95% CI: 53.0% - 67.5%).This means that YiDiXie ™ -SS has extremely high sensitivity and relatively high specificity in prostate tumors.

**Table 2.** The performance of YiDiXie™ - SS.

|  | Malignant | Benign | Total |
| --- | --- | --- | --- |
|  | 292 | 172 | 464 |
| Positive | 292 | 68 | 360 |
| Negative | 0 | 104 | 104 |

SEN, Sensitivity. SPE, Specificity. Two-sided 95% Wilson confidence intervals were calculated.

### Diagnostic performance of YiDiXie™-HS

As shown in Table 3, the sensitivity of YiDiXie™ -HS was 93.8% (95% CI: 90.5% - 96.1%) and its specificity was 86.0% (95% CI: 80.1% - 90.4%). This means that YiDiXie ™ -HS has high sensitivity and specificity in prostate tumors.

**Table 3.** Performance of YiDiXie™ -HS.

|  | Malignant | Benign | Total |
| --- | --- | --- | --- |
|  | 292 | 172 | 464 |
| Positive | 274 | 24 | 298 |
| Negative | 18 | 148 | 166 |

SEN, Sensitivity. SPE, Specificity. Two-sided 95% Wilson confidence intervals were calculated.

### Diagnostic performance of YiDiXie™-D

As shown in Table 4, the sensitivity of YiDiXie™ -D was 80.8% (95% CI: 75.9% - 84.9%) and its specificity was 93.0% (95% CI: 88.2% - 96.0%). This means that YiDiXie ™ -D has relatively high sensitivity and very high specificity in prostate tumors.

**Table 4.**
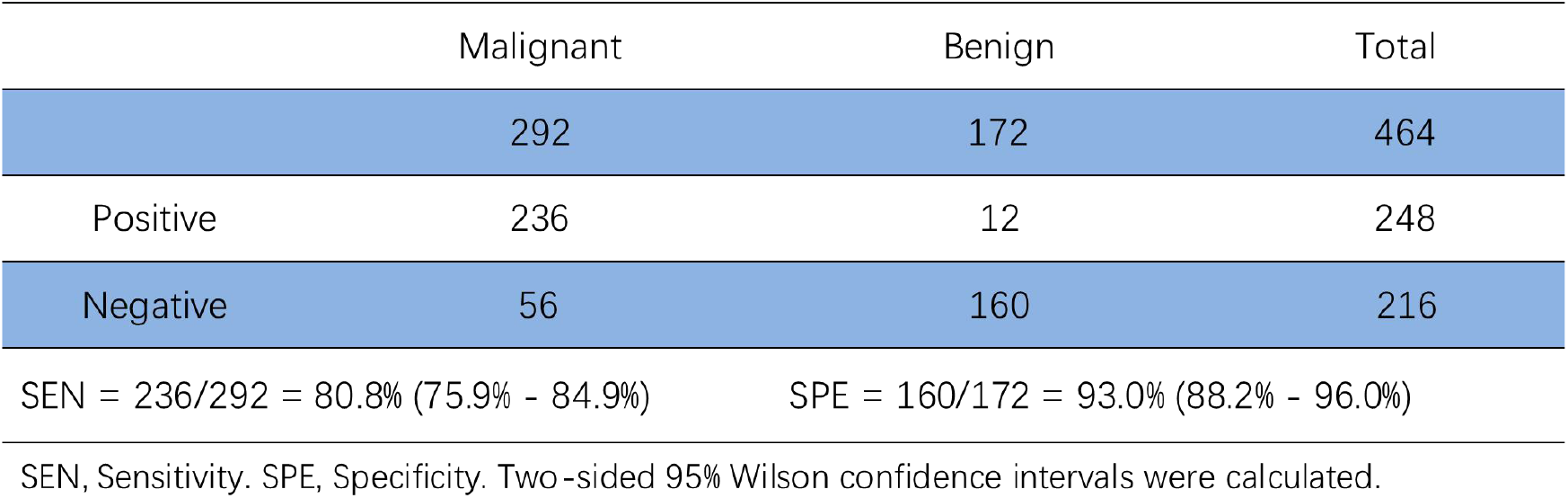
Performance of YiDiXie™ -D.

|  | Malignant | Benign | Total |
| --- | --- | --- | --- |
|  | 292 | 172 | 464 |
| Positive | 236 | 12 | 248 |
| Negative | 56 | 160 | 216 |

SEN = 236/292 = 80.8% (75.9% - 84.9%)
SPE = 160/172 = 93.0% (88.2% - 96.0%)
SEN, Sensitivity. SPE, Specificity. Two-sided 95% Wilson confidence intervals were calculated.

### Diagnostic performance of YiDiXie™-SS in PSA-positive patients

To address the challenge of high PSA false-positive rate, YiDiXie ™ -SS was applied to PSA-positive patients.

As shown in Table 5, the sensitivity of YiDiXie™ -SS in PSA-positive patients was 100% (95% CI: 98.6% - 100%), and the specificity was 57.7% (95% CI: 49.8% - 65.2%). This means that the application of YiDiXie™-SS reduces the PSA false-positive rate by 57.7% (95% CI: 49.8% - 65.2%) with essentially no increase in malignancy leakage.

**Table 5.** Performance of different Items.

| Items | Total N | Positive | Sensitivity % (95% CI)<sup>a</sup> | Total N | Negative | Specificity % (95% CI)<sup>b</sup> |
| --- | --- | --- | --- | --- | --- | --- |
| YiDiXie™-SS in PSA-positive | 279 | 279 | 100% (98.6% - 100%) | 156 | 90 | 57.7% (49.8% - 65.2%) |
| YiDiXie™-HS in PSA-negative | 13 | 12 | 92.3% (66.7% - 99.6%) | 16 | 13 | 81.3% (57.0% - 93.4%) |
| YiDiXie™-D in PSA-positive | 279 | 226 | 81.0% (76.0% - 85.2%) | 156 | 145 | 92.9% (87.8% - 96.0%) |
| YiDiXie™-D in PSA-negative | 13 | 10 | 76.9% (49.7% - 91.8%) | 16 | 15 | 93.8% (71.7% - 99.7%) |
| YiDiXie™-SS in MRI-positive | 159 | 159 | 100% (97.6% - 100%) | 94 | 56 | 59.6% (49.5% - 68.9%) |
| YiDiXie™-HS in MRI-negative | 51 | 46 | 90.2% (79.0% - 95.7%) | 12 | 10 | 83.3% (55.2% - 97.0%) |
| YiDiXie™-D in MRI-positive | 159 | 133 | 83.6% (77.1% - 88.6%) | 94 | 87 | 92.6% (85.4% - 96.3%) |
| YiDiXie™-D in MRI-negative | 51 | 39 | 76.5% (63.2% - 86.0%) | 12 | 10 | 83.3% (55.2% - 97.0%) |

CI, confidence interval. <sup>a,b</sup> Two-sided 95% Wilson confidence intervals were calculated.

### Diagnostic performance of YiDiXie™-HS in PSA-negative patients

To solve the challenge of high PSA false-negative rate, YiDiXie ™ -HS was applied to PSA-negative patients.

As shown in Table 5, the sensitivity of YiDiXie™ -HS in PSA-negative patients was 92.3% (95% CI: 66.7% - 99.6%), and the specificity was 81.3% (95% CI: 57.0% - 93.4%). This means that the application of YiDiXie ™-HS reduced the PSA false negative rate by 92.3% (95% CI: 66.7% - 99.6%).

### Diagnostic performance of YiDiXie™-D in PSA-positive patients

To further reduce the rate of PSA false positives, YiDiXie ™-D, which has a relatively high sensitivity and very high specificity, was therefore applied.

As shown in Table 5, YiDiXie ™ -D had a sensitivity of 81.0% (95% CI: 76.0% - 85.2%) and its specificity was 92.9% (95% CI: 87.8% - 96.0%) in PSA-positive patients. This means that YiDiXie™-D reduces the PSA false positive rate by 92.9% (95% CI: 87.8% - 96.0%)..

### Diagnostic performance of YiDiXie™-D in PSA-negative patients

In order to reduce the PSA false-negative rate while maintaining high specificity, YiDiXie ™ -D, which has relatively high sensitivity and very high specificity, was applied.

As shown in Table 5, YiDiXie ™ -D has a sensitivity of 76.9% (95% CI: 49.7% - 91.8%) in PSA-negative patients and its specificity is 93.8% (95% CI: 71.7% - 99.7%). This means that YiDiXie™-D reduces the PSA false-negative rate by 76.9% (95% CI: 49.7% - 91.8%) while maintaining high specificity.

### Diagnostic performance of YiDiXie™-SS in enhanced MRI-positive patients

To address the challenge of high false-positive rate of enhanced MRI, YiDiXie™-SS was applied to enhanced MRI-positive patients.

As shown in Table 5, YiDiXie ™ -SS had a sensitivity of 100% (95% CI: 97.6% - 100%) and a specificity of 59.6% (95% CI: 49.5% - 68.9%) in enhanced MRI-positive patients. This means that the application of YiDiXie ™ -SS reduces the false-positive rate of enhanced MRI by 59.6% (95% CI: 49.5% - 68.9%) with essentially no increase in malignancy leakage.

### Diagnostic performance of YiDiXie™-HS in enhanced MRI-negative patients

In order to solve the problem of high false-negative rate of enhanced MRI, YiDiXie™-HS was applied to enhanced MRI-negative patients.

As shown in Table 5, the sensitivity of YiDiXie™ -HS in enhanced MRI-negative patients was 90.2% (95% CI: 79.0% - 95.7%), and the specificity was 83.3% (95% CI: 55.2% - 97.0%). This means that the application of YiDiXie ™ -HS reduced the false negative rate of enhanced MRI by 90.2% (95% CI: 79.0% - 95.7%).

### Diagnostic performance of YiDiXie™-D in enhanced MRI-positive patients

To further reduce the rate of enhanced MRI false positives, YiDiXie ™-D, which has a relatively high sensitivity and very high specificity, was therefore applied.

As shown in Table 5, YiDiXie ™ -D had a sensitivity of 83.6% (95% CI: 77.1% - 88.6%) and its specificity was 92.6% (95% CI: 85.4% - 96.3%) in enhanced MRI-positive patients. This means that YiDiXie ™ -D reduces the enhanced MRI false positive rate by 92.6% (95% CI: 85.4% - 96.3%).

### Diagnostic performance of YiDiXie™-D in enhanced MRI-negative patients

In order to reduce the enhanced MRI false-negative rate while maintaining high specificity, YiDiXie ™ -D, which has relatively high sensitivity and very high specificity, was applied.

As shown in Table 5, YiDiXie ™ -D has a sensitivity of 76.5% (95% CI: 63.2% - 86.0%) in enhanced MRI-negative patients and its specificity is 83.3% (95% CI: 55.2% - 97.0%). This means that YiDiXie ™ -D reduces the enhanced MRI false-negative rate by 76.5% (95% CI: 63.2% - 86.0%) while maintaining high specificity.

## DISCUSSION

### Clinical significance of YiDiXie™-SS in PSA or enhanced MRI-positive patients

YiDiXie ™ tests include three distinct tests: YiDiXie ™ -HS, YiDiXie ™ -SS, and YiDiXie ™ -D. Among them, YiDiXie ™ -HS combines high sensitivity and high specificity. YiDiXie ™ -SS has extremely high sensitivity for all malignant tumor types, but slightly lower specificity. YiDiXie ™-D is extremely high specific for all forms of malignant tumors but has lesser sensitivity.

The sensitivity and specificity of further diagnostic techniques are crucial in PSA or enhanced MRI-positive patients. On the one hand, sensitivity is important. A higher percentage of false negatives results from lower sensitivity. When this diagnostic approach yields a negative result, the patient’s diagnosis is usually terminated. A larger false-negative rate means that more cancerous tumors are overlooked, which will lead to delays in their treatment, progression of the malignancy, and possibly even development of advanced stages. Patients will thus have to bear the adverse consequences of poor prognosis, poor quality of life and high treatment costs.

On the other hand, specificity is important. Lower specificity means a higher rate of false positives. When the results of this diagnostic method are positive, the diagnosis of prostate cancer is usually confirmed by a puncture biopsy. A higher false-positive rate means that more non-prostate cancer cases undergo puncture biopsy. This undoubtedly substantially increases the patient’s mental anguish, cost of testing, physical harm and other adverse consequences^11,12^.

Consequently, there is a trade-off between “fewer malignant tumors missed” and “fewer benign tumors misdiagnosed” when it comes to sensitivity and specificity. When non-prostate cancer cases is mistakenly identified as malignant tumor, aspiration biopsy is typically performed instead of surgical resection. Therefore, PSA false positives do not lead to serious consequences in terms of organ loss. Thus, for PSA-positive patients, “fewer missed diagnoses of malignant tumors” is considerably more essential than “fewer misdiagnoses of benign tumors.” Therefore, YiDiXie ™ -SS was chosen for reducing the false-positive rates of PSA rather than YiDiXie™-HS or YiDiXie™ -D.

As shown in Tables 5, YiDiXie ™ -SS had a sensitivity of 100% (95% CI: 98.6% - 100%),100% (95% CI: 97.6% - 100%) and a specificity of 57.7% (95% CI: 49.8% - 65.2%), 59.6% (95% CI: 49.5% - 68.9%). These results indicate that YiDiXie ™ -SS reduces false positives by 57.7% (95% CI: 49.8% - 65.2%), 59.6% (95% CI: 49.5% - 68.9%) for PSA and enhanced MRI, respectively, while maintaining a sensitivity of nearly 100%.

As mentioned earlier, missed diagnosis of prostate cancer means delayed treatment, and false-positive PSA or enhanced MRI results mean wrong puncture biopsy. These results imply that, with essentially no increase in malignancy leakage, YiDiXie ™ -SS substantially reduces the probability of erroneous puncture biopsies in PSA or enhanced MRI-positive patients.

In other words, YiDiXie ™ -SS considerably decreases the emotional suffering, expensive examination costs, physical injuries, and other harmful consequences for PSA or enhanced MRI-false positive patients without significantly increasing the delay in the treatment of malignant tumors. Therefore, YiDiXie ™-SS meets the clinical needs well and has important clinical significance and wide application prospects.

### Clinical significance of YiDiXie™-HS in PSA or enhanced MRI-negative patients

For PSA or enhanced MRI-negative patients, it is important that further diagnostic methods have both sensitivity and specificity. The higher false-negative rate implies that more prostate cancers are underdiagnosed. The higher false-positive rate implies that more benign prostate cases are misdiagnosed. Usually, patients with benign prostate cases misdiagnosed as prostate cancer usually undergo puncture biopsy, which is neither radical surgery nor does it affect the patient’s prognosis, and the cost of its puncture biopsy is much lower than the cost of treatment for advanced cancer. Therefore, for PSA or enhanced MRI-negative patients, the “ harm of misdiagnosis of benign prostate cases” is lower than the “harm of underdiagnosis of prostate cancer”. Furthermore, the negative predictive value was higher in PSA or enhanced MRI-negative patients. As a result, both higher false-positive and false-negative rates can lead to significant harms. Therefore, YiDiXie ™-HS with high sensitivity and specificity was chosen to reduce the PSA or enhanced MRI false-negative rate.

As shown in Tables 5, YiDiXie ™ -HS had a sensitivity of 92.3% (95% CI: 66.7% - 99.6%), 90.2% (95% CI: 79.0% - 95.7%), and specificity of 81.3% in PSA and enhanced MRI-negative patients 81.3% (95% CI: 57.0% - 93.4%)?83.3% (95% CI: 55.2% - 97.0%). This means that YiDiXie ™ -HS reduced PSA and enhanced MRI false negatives by 92.3% (95% CI: 66.7% - 99.6%), 90.2% (95% CI: 79.0% - 95.7%), respectively.

The above results imply that YiDiXie ™ -HS substantially reduces the probability of false-negative miss-diagnosis of malignant tumors by PSA or enhanced MRI. Therefore, YiDiXie™-HS meets the clinical needs well and has important clinical significance and wide application prospects.

### Clinical significance of YiDiXie™-D

For patients with prostate tumors, YiDiXie™-D, with its relatively high sensitivity and very high specificity, can be used to further reduce the rate of false positives or significantly reduce the rate of false negatives on PSA or enhanced MRI while maintaining high specificity.

As shown in Table 5, the sensitivity of YiDiXie™ -D in patients with positive PSA or enhanced MRI was 81.0% (95% CI: 76.0% - 85.2%), 83.6% (95% CI: 77.1% - 88.6%), and its specificity was 92.9% (95% CI: 87.8% - 96.0%), 92.6% (95% CI : 85.4% - 96.3%);The sensitivity of YiDiXie™-D in patients with negative PSA or enhanced MRI was 76.9% (95% CI: 49.7% - 91.8%), 76.5% (95% CI: 63.2% - 86.0%), and its specificity was 93.8% (95% CI: 71.7% - 99.7%), 83.3% (95% CI: 55.2% - 97.0%).These results indicate that YiDiXie ™ -D reduced PSA or enhanced MRI false positives by 92.9% (95% CI: 87.8% - 96.0%), 92.6% (95% CI: 85.4% - 96.3%) or, while maintaining a high level of specificity, by 76.9% (95% CI: 49.7% - 91.8%), 76.5% (95% CI: 63.2% - 86.0%), respectively.

The above results imply that YiDiXie ™ -D substantially reduces the probability of wrong puncture biopsy in prostate tumors. Therefore, YiDiXie™-D well meets the clinical needs and has important clinical significance and wide application prospects.

### YiDiXie™ tests are expected to solve 2 challenges in prostate cancer

Firstly, YiDiXie ™ -SS significantly reduces the risk of misdiagnosis of benign prostate cases as prostate cancer. On the one hand, YiDiXie ™ -SS substantially reduces the probability of incorrect puncture biopsy in benign prostate cases with essentially no increase in missed prostate cancer diagnosis. As shown in Tables 5, YiDiXie ™ -SS reduced PSA and enhanced MRI false positives by 57.7% (95% CI: 49.8% - 65.2%), and 59.6% (95% CI: 49.5% - 68.9%), respectively, with essentially no increase in prostate cancer missed diagnosis. Thus, YiDiXie ™ -SS significantly reduces the range of adverse outcomes associated with unnecessary prostate puncture biopsy with essentially no increase in delayed treatment of prostate cancer.

On the other hand, YiDiXie ™ -SS reduces physician burdens and allows for rapid treatment of malignancies that might otherwise be postponed. When PSA or enhanced MRI result is positive, the patient is often treated with a puncture biopsy^10^. The number of physician determines how many biopsies are performed. That appointments for prostate biopsies can take months or even more than a year are widely available around the world. It is inevitable that the treatment of malignant cases would be delayed. It is not uncommon for PSAor enhanced MRI-positive patients awaiting treatment to develop malignant progression or even distant metastases. As shown in Tables 5, YiDiXie ™ -SS reduced PSA and enhanced MRI false positives by 57.7% (95% CI: 49.8% - 65.2%), and 59.6% (95% CI: 49.5% - 68.9%), respectively, with essentially no increase in missed prostate cancer diagnoses. As a result, YiDiXie™-SS greatly relieves physicians from unnecessary workloads and facilitates the timely treatment of malignant tumor cases that would otherwise be delayed.

Secondly, YiDiXie™-HS greatly reduces the risk of missed prostate cancer diagnosis.When PSA or enhanced MRI is negative, prostate cancer is usually ruled out temporarily. The high rate of false-negative PSA or enhanced MRI results in delayed treatment for a large number of prostate cancer patients. As shown in Tables 5, YiDiXie™-HS reduced PSA and enhanced MRI false-negative margins by 92.3% (95% CI: 66.7% - 99.6%), 90.2% (95% CI: 79.0% - 95.7%), respectively. Thus, YiDiXie™-HS substantially reduces the probability of false-negative PSA or enhanced MRI miss-diagnosis of malignant tumors and facilitates timely diagnosis and treatment of prostate cancer patients who would otherwise be delayed in treatment.

Thirdly, YiDiXie ™ -D is expected to further address the challenges of “high false positive rate” and “high false negative rate”. As shown in Table 5, YiDiXie ™ -D reduced PSA or enhanced MRI false positives by 92.9% (95% CI: 87.8% - 96.0%) and 92.6% (95% CI: 85.4% - 96.3%), respectively, or 76.9% (95% CI: 49.7% - 91.8%), 76.5% (95% CI: 63.2% - 86.0%) for the reduction of false negatives on PSA or enhanced MRI, respectively, while maintaining a high level of specificity. Thus, YiDiXie ™ -D further reduced the risk of puncture biopsy in prostate tumors.

Finally, YiDiXie ™ tests offer “just-in-time diagnosis” for PSA or enhanced MRI-positive patients. On the one hand, YiDiXie ™ tests are non-invasive and only requires a small amount of blood, allowing patients to complete the diagnostic process at home. The YiDiXie™ test requires only 20 microliters of serum, equivalent to around 1 drop of whole blood (1 drop of whole blood is approximately 50 microliters, which yields 20-25 microliters of serum). Considering the pre-test sample quality assessment test and 2-3 repetitions, 0.2 ml of whole blood is sufficient for YiDiXie ™ tests. General patients can obtain the 0.2 ml of finger blood at home with a finger blood collection needle, without the need for venous blood collection by medical staff. Patients can complete the entire diagnostic process non-invasively without leaving their homes.

On the other hand, YiDiXie™ tests have nearly infinite diagnostic capability. Figure 1 shows the basic flow chart of YiDiXie™ tests. This means that YiDiXie™ tests not only don’t require a doctor or medical equipment, but also does not require medical staff to collect blood. As a result, YiDiXie™ tests have an almost infinite diagnostic capacity and is totally independent of the quantity of clinician and medical facilities. Thus, YiDiXie™ tests enable patients with prostate tumor to receive a “just-in-time diagnosis” without making them wait anxiously for an appointment.

**Figure 1.**
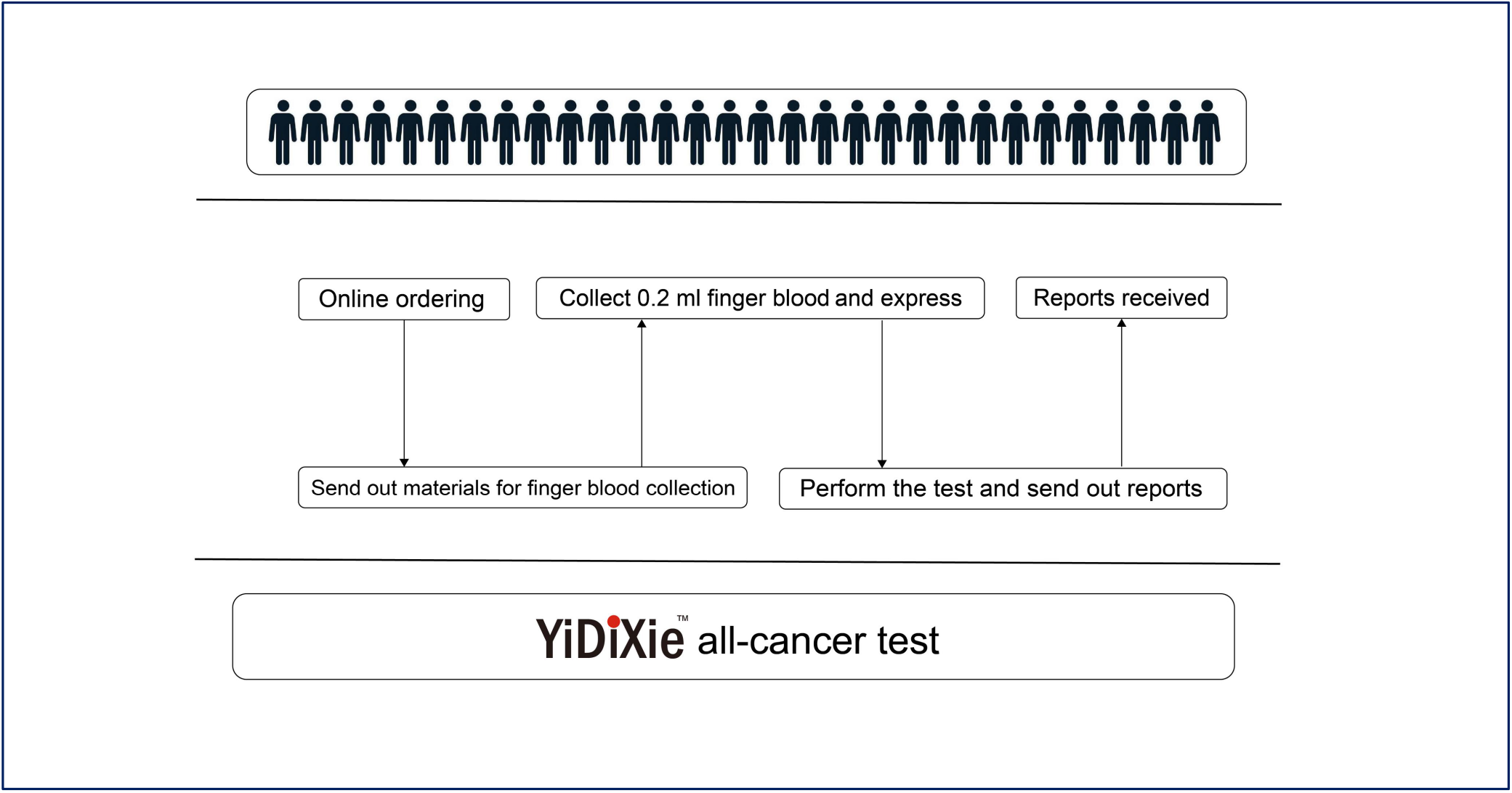
Basic flowchart of YiDiXie™ tests.

In short, YiDiXie™ tests can play an important role in PSA or enhanced MRI-positive patients, and are expected to solve the problem of “high false-positive rate” and “high false-negative rate” of PSA or enhanced MRI.

### Limitations of the study

First, the number of cases in this study was small, and future studies with larger sample sizes are needed for further evaluation.

Second, this study was a case-control study of hospitalized patients, and future cohort studies of normal population are needed for further evaluation.

Final, this study was a single-center study, which may have led to some degree of bias. Future multi-center studies are needed for further evaluation.

## CONCLUSION

YiDiXie ™ -SS has very high sensitivity and relatively high specificity in prostate tumors.YiDiXie ™ -HS has high sensitivity and high specificity in prostate tumors.YiDiXie ™ -D has relatively high sensitivity and very high specificity in prostate tumors. YiDiXie™-SS significantly reduces the rate of PSA or enhanced MRI false positives with essentially no increase in delayed treatment for prostate cancer.YiDiXie ™ -HS significantly reduces the rate of PSA or enhanced MRI false negatives.YiDiXie™-D significantly reduces the rate of PSA or enhanced MRI false positives or significantly reduces the rate of false negatives thereof while maintaining a high level of specificity. YiDiXie ™ tests can play an important role in prostate cancer, and are expected to solve the problem of “high false-positive rate” and “high false-negative rate” of PSA or enhanced MRI.

## Data Availability

All data produced in the present study are contained in the manuscript.

